# Neonatal-onset autoinflammation and immunodeficiency caused by heterozygous missense mutation of the proteasome subunit β-type 9

**DOI:** 10.1101/2021.02.01.21250077

**Authors:** Nobuo Kanazawa, Hiroaki Hemmi, Noriko Kinjo, Hidenori Ohnishi, Jun Hamazaki, Hiroyuki Mishima, Akira Kinoshita, Tsunehiro Mizushima, Satoru Hamada, Kazuya Hamada, Norio Kawamoto, Saori Kadowaki, Yoshitaka Honda, Kazushi Izawa, Ryuta Nishikomori, Miyuki Tsumura, Yusuke Yamashita, Shinobu Tamura, Takashi Orimo, Toshiya Ozasa, Takashi Kato, Izumi Sasaki, Yuri Fukuda-Ohta, Naoko Wakaki-Nishiyama, Yutaka Inaba, Kayo Kunimoto, Satoshi Okada, Takeshi Taketani, Koichi Nakanishi, Shigeo Murata, Koh-ichiro Yoshiura, Tsuneyasu Kaisho

**Author notes:** **Corresponding authors** Nobuo Kanazawa MD, PhD, Department of Dermatology, Hyogo College of Medicine, 1-1 Mukogawa-cho, Nishinomiya, Hyogo 663-8501, Japan, Tsuneyasu Kaisho MD, PhD, Department of Immunology, Institute of Advanced Medicine, Wakayama Medical University, 811-1 Kimiidera, Wakayama, Wakayama 641-8509, Japan. These authors contributed equally to this work (Nobuo Kanazawa, Hiroaki Hemmi).

## Abstract

**BACKGROUND:** Defective proteasome activities due to genetic mutations lead to an autoinflammatory disease, termed as proteasome-associated autoinflammatory syndromes (PRAAS). In PRAAS relapsing inflammations and progressive wasting are common, but immunodeficiency has not been reported.

**METHODS:** We studied two unrelated Japanese infants with PRAAS-like manifestations. We have also generated and analyzed the mice carrying the candidate mutation found in the patients.

**RESULTS:** Both patients showed neonatal-onset skin rash, myositis and basal ganglia calcification, similar to PRAAS patients. Meanwhile, they manifested distinct phenotypes, including pulmonary hypertension and immunodeficiency without lipoatrophy. We identified a novel *de novo* heterozygous missense mutation, G156D, in a proteasome subunit gene, *PSMB9*, encoding β1i, in the two patients. Maturation and activity of the immunoproteasome were impaired, but ubiquitin accumulation was hardly detected not only in patient-derived cells and samples but also in *Psmb9*^*G156D/+*^ mice. As an immunodeficient phenotype, one patient showed decrease of B cells and increase of monocytes, while the other patient showed decrease of CD8 T cells. The proteasome defects and immunodeficient phenotypes were recapitulated in *Psmb9*^*G156D/+*^ mice.

**CONCLUSIONS:** The PSMB9 G156D is a unique mutation in proteasome subunits in causing defects by its heterozygosity, affecting two β rings interaction and leading to immunodeficiency. The mutant mice are the first mice model for analyzing proteasome dysfunctions in PRAAS. We here propose the term, proteasome-associated autoinflammation and immunodeficiency disease (PRAID), as an umbrella name for our cases, PRAAS with immunodeficiency, as well as PRAAS described so far.

## INTRODUCTION

Mutations of *proteasome subunit β-type 8 (PSMB8)* were first detected in autoinflammatory diseases characterized by systemic relapsing inflammations and progressive wasting, such as Nakajo-Nishimura syndrome and chronic atypical neutrophilic dermatosis with elevated temperature (CANDLE).^1-4^ Subsequently, identification of mostly biallelic mutations of other proteasome subunit and chaperone genes, leading to loss-of-function of the proteasome, has defined a new disease entity as proteasome-associated autoinflammatory syndrome (PRAAS).^5-8^

Proteasome is a protein complex involved in intracellular protein homeostasis by degrading unnecessary or useless proteins tagged with polyubiquitin.^9,10^ All eukaryotic cells have the constitutive 26S proteasome which consists of a 20S core particle and two 19S regulatory particles. 20S core complex is composed of α1-α7 and β1-β7 subunits, among which β1, β2, and β5 mediate the protease activity. In hematopoietic cells or fibroblasts stimulated with cytokines, inducible subunits, β1i (coded by *PSMB9*), β2i (coded by *PSMB10*), and β5i (coded by *PSMB8*), are substituted for β1, β2, and β5, respectively, to form immunoproteasome. Especially, in thymic cortical epithelial cells, β5t (coded by *PSMB11*), instead of β5i, as well as β1i and β2i are expressed to form thymoproteasome. Immunoproteasome and thymoproteasome are involved not only in protein degradation but also in generation of antigen peptides presented with major histocompatibility complex (MHC) class I molecules and CD8 T cell repertoire, population and responses.

Here we identified a novel *de novo PSMB9* heterozygous missense mutation, G156D, in two unrelated Japanese patients with manifestations, including autoinflammation and immunodeficiency, which were similar to, but distinct from those of PRAAS patients. The proteasome defect and immunodeficient phenotypes were recapitulated in *Psmb9*^*G156D/+*^ mice.

## METHODS

### Patients

Written informed consent was obtained from the patients’ parents and all other study participants.

### Genetic studies

Whole-exome sequencing was performed by using genomic DNAs obtained from peripheral blood of patients and parents as described previously.^11^ Identified candidate mutations were confirmed by Sanger sequencing. T-cell receptor recombination excision circles (TREC) and immunoglobulin (Ig) κ-deleting recombination excision circles (KREC) levels in the patients’ peripheral blood mononuclear cells (PBMC) were measured by real-time polymerase chain reaction (PCR) as described previously.^12^ Interferon (IFN) score was determined as described previously.^13,14^ Details are described in the Supplementary Appendix.

### Structural and protein analyses

Structural modeling of wildtype and mutated immunoproteasomes and proteasome analysis were performed as described previously.^3^ Immunohistochemistry was performed with antibodies (Abs) against phosphorylated signal transducers and activators of transcription 1 at Tyr701 (p-STAT1) or ubiquitin. Details are described in the Supplementary Appendix.

### Generation and analysis of *Psmb9 G156D* mutant mice

Mutant mice were generated by the CRISPR/Cas9 method and analyzed. Details are described in the Supplementary Appendix.

## RESULTS

### Clinical phenotype

Two patients from nonconsanguineous families were enrolled in this study. These patients showed the common features, such as neonatal-onset fever, increased inflammatory reactants, skin rash, myositis, liver dysfunction, pulmonary arterial hypertension and basal ganglia calcification. Autoantibodies associated with dermatomyositis were not detected in either patient. The serum inflammatory cytokine levels, such as IL-6, IL-18, and IP-10, were increased in both patients. In patient 2, IFN-α was not only increased in the serum but also detected in central spinal fluid. Furthermore, the IFN score was positive in the whole blood and STAT1 phosphorylation was enhanced in the skin lesion and dermal fibroblasts (Fig. S2C). Patient 2 also showed manifestations with hemophagocytic lymphohistiocytosis.^15^ Concerning immunological functions, patient 1 showed decrease of serum IgG level and B cell numbers (Table S1), and required IgG supplementation to control manifestations. Patient 1 also exhibited low levels of both TREC and KREC, indicating T as well as B cell defects. Meanwhile, patient 2 showed decrease of monocytes, CD8 T and γδ T cells and almost absence and decreased activity of natural killer (NK) cells (Table S1). Interestingly, polyoma (BK/JC) viruses were detected in both patient’s peripheral bloods (Table 1).

**Table 1.** Comparison of PRAAS, patients 1, patient 2 and *Psmb9*^*G156D/+*^ mice.

|  |  | **PRAAS** | **Patient 1** | **Patient 2** | ***Psmb9*<sup>G156D/+</sup> mice** |
| --- | --- | --- | --- | --- | --- |
| **Clinical findings** | Fever | + | + | + | - |
|  | Skin rash | + | + | + | - |
|  | Myositis | + | + | + | NA |
|  | Liver dysfunction | + | +(fibrosis) | + | NA |
|  | Pneumonia | + | + | + | NA |
|  | Splenomegaly | + | + | + | - |
|  | Lipoatrophy | + | - | - | - |
|  | Pulmonary hypertension | - | + | + | NA |
|  | Basal ganglia calcification | + | + | + | NA |
|  | IFN score | Positive | - | Positive | - |
|  | Viremia | - | Polyoma virus + | Polyoma virus + | NA |
| **Blood analysis** | Neutrophil | Variable | ↑ | → | ↑ |
|  | Monocyte | Variable | ↑ | ↓ | ↑ |
|  | NK cell | Variable | → | ↓ | → |
|  | Dendritic cell | NA | NA | NA | ↓ |
|  | T cell | Variable | →(CD4) ↑(CD8) ↓(γδ) | ↓(CD8) ↓(γδ) | ↓ |
|  | B cell | Variable | ↓ | → | ↓ |
|  | Serum Ig | ↑ | ↓(IgG) | Unevaluable | ↓ |
| **Proteasome analysis** | β1i maturation | Variable | Defect | Defect | Defect |
|  | Proteasome activity | 20S, 26S defect | 20S defect | 20S defect | 20S defect |
|  | Ubiquitin accumulation | ++ | +/- | +/- | - |

NA, not assessed.

The detailed case reports were described in Supplementary Appendix and summarized in Table 1. In summary, these two patients initially showed mainly severe autoinflammatory phenotypes and later manifested the immunodeficiency phenotype with periodic inflammatory exacerbation. Clinical features of both patients were partially overlapped with PRAAS. However, unlike PRAAS, they apparently lacked lipoatrophy, one of the main characteristics of PRAAS, and showed pulmonary hypertension. More importantly, they also manifested combined immunodeficiency (at least B and T cells in patient 1 and T and NK cells in patient 2), which is not seen in PRAAS (Table 1).

### Genetic and functional analysis

In Patient 1, no pathogenic mutation was first identified in *MEFV, MVK, NLRP3, NOD2, TNFRSF1A* or *PSMB8*. Then, a panel of ubiquitin-proteasome-system, autophagy and interferonopathy-related genes (Table S2A) were sequenced and two heterozygous *PSMB9* (NM_002800) c.467G>A (p.G156D) and *PSMD9* (NM_002813) c.31G>A (p.G11S) mutations, which could be pathogenic, were identified. The former is a novel *de novo* mutation in the immunoproteasome-specific β1i subunit gene and 156G is well conserved among β1 and β1i across species (Fig. 1A). The latter, a rare polymorphism (rs183923514), was also detected in the patient’s relative without any autoinflammatory manifestations and 11G is not conserved even in mouse. Furthermore, whole exome sequencing of genomic DNAs from the patient and the parents revealed two homozygous, 1 compound heterozygous and 29 *de novo* mutations in patient 1 (Table S3). Among these genes, only *PSMB9* was associated with inflammation. In patient 2, a panel of autoinflammatory disease-causing genes (Table S2B) were analyzed and *de novo PSMB9* c.467G>A (p.G156D) heterozygous mutation, i.e. the same mutation found in patient1, was identified. Whole exome sequencing of genomic DNAs from the patient and the parents identified two homozygous, 1 compound heterozygous and 12 *de novo* mutations in patient 2 (Table S4). Among these genes, only *PSMB9* was associated with inflammation. Structural modeling revealed that G156 in β1i is located on the interface between two β rings and G156D substitution affects their interaction (Fig. 1B), while keeping the active site conformation (Fig.S1A). Consistent with this modeling, immunoblotting analysis of patient 1-derived immortalized B cells showed increase of immunoproteasome intermediate containing Ump1 and immature β1i and severely diminished incorporation of all induced subunits into the 20S complex (Figs. 1C and S1B). Meanwhile, in the 26S proteasome of patient 1-derived cells, β2i and β5i incorporation was comparable to that of patient 1’s relative-derived control cells, although β1i incorporation was impaired and immature β1i was detected. In patient 1-derived cells, the catalytic activity of the 20S, but not the 26S proteasome, was severely impaired, whereas both the 20S and 26S proteasome showed severe defects in the activities in PRAAS patient-derived cells (Fig. 1D). Ubiquitin accumulation, which is caused by the 26S proteasome defect and is common in PRAAS, was unremarkable in the patients’ skin lesion (Figs. 1E and S2D). In patient 2, unstimulated fibroblasts showed no significant defects in components of the proteasomes (Fig. S2A). However, IFNγ-stimulated fibroblasts showed defects in maturation of β1i and β5i and incorporation of β1i into the 20S and 26S proteasome and impaired catalytic activity of the 20S complex, with normal 26S proteasome activity (Fig. S2A, B).

**Figure 1.**
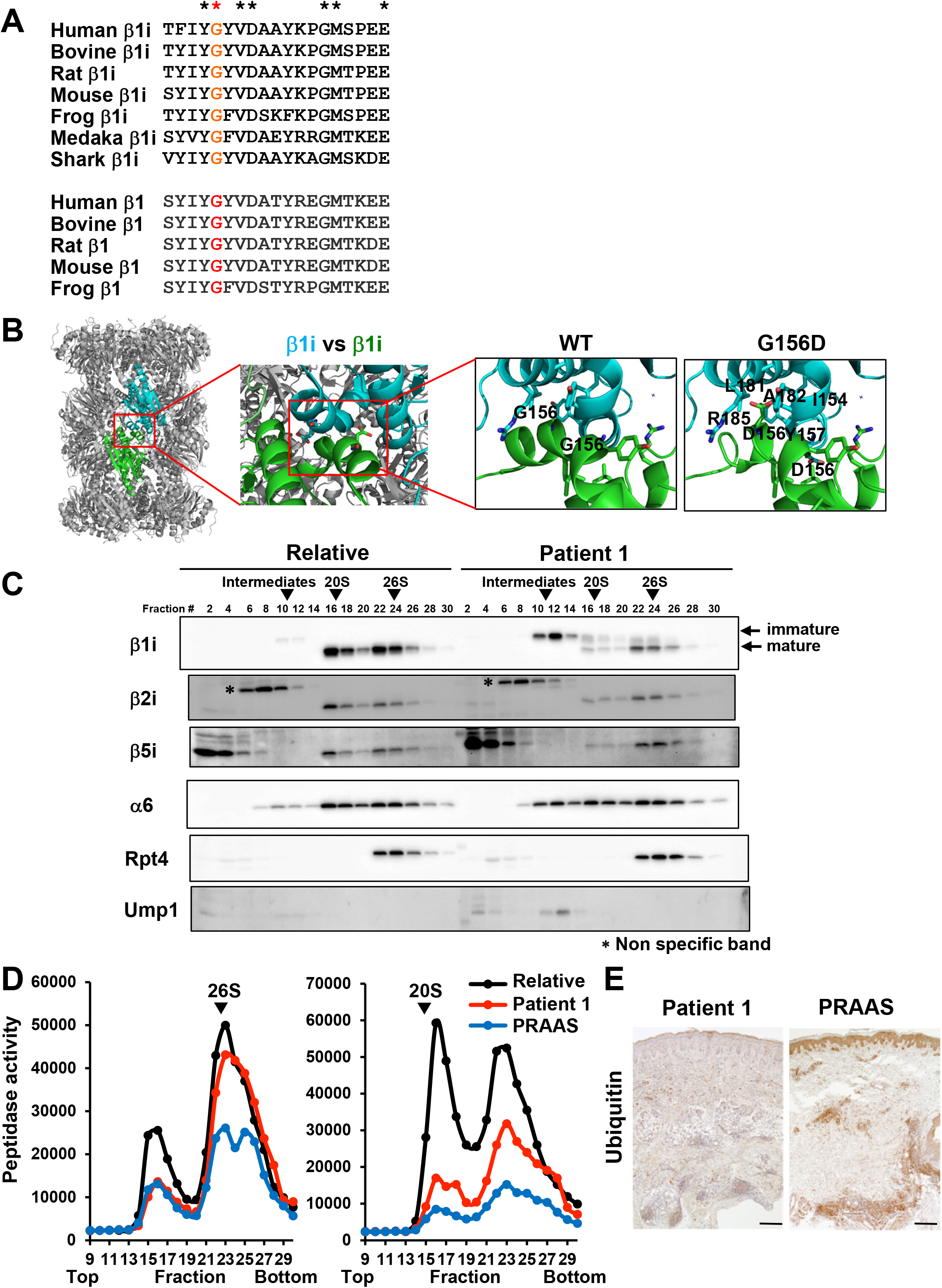
Genetic and protein analysis of the patients. Panel A shows multiple alignment of PSMB9 (β1i) and PSMB6 (β1) in various species and high conservation of G156 in murine PSMB9 (red). Panel B shows structures of wild type and PSMB9 G156D mutant of 20S proteasome. Structural models of PSMB9 G156D was created from the β1i-subunit structure [Protein Data Bank (PDB) ID code 3UNH]. Overall structure of 20S is shown as a ribbon model. β1i subunits are shown as green and cyan. D156 and some of the potential interacting residues of PSMB9 G156D mutant are shown in stick representation (green and cyan). Panels C and D shows the proteasome analysis of immortalized B cells from patient 1 and the patient’s relative. Cell extracts were prepared and fractionated by glycerol gradient centrifugation. Panel C shows immunoblot analysis of each fraction using Abs against the indicated proteins. Panel D shows chymotrypsin-like activity of each fraction, which was measured by using Suc-LLVY-AMC as a substrate in the absence (left) or presence (right) of 0.0025% SDS. Panel E shows the skin biopsy samples of patient 1 and a PRAAS patient stained with anti-ubiquitin Ab. Scale bars represent 100 μm.

### Analysis of mice carrying the PSMB9 G156D mutation

In order to clarify how the PSMB9 G156D mutation contributes to the patients’ manifestations, we generated mice carrying the mutation by the CRISPR/Cas9 method. *Psmb9*^*G156D/G156D*^ mice died within 6-7 months old, while survival rate of *Psmb9*^*G156D/+*^ mice was similar to that of wildtype mice (Fig. S3A). *Psmb9*^*G156D/+*^ mice appeared healthy at glance and lacked a sign of lipoatrophy or inflammation (Fig. S3B, Table 1). Components and catalytic activities of proteasomes in unstimulated embryonic fibroblasts were comparable among *Psmb9*^*+/+*^, *Psmb9*^*G156D/+*^ and *Psmb9*^*G156D/G156D*^ mice (Fig. S3C, D). However, in IFNγ-stimulated embryonic fibroblasts, immature β1i and β2i, which were undetectable in *Psmb9*^*+/+*^ mice, were observed and incorporation of β1i into the 20S complex was severely impaired, although incorporation of β1i into the 26S proteasome was mildly defective in *Psmb9*^*G156D/+*^ mice (Fig. 2A). Neither β1i nor β2i were hardly detected in 20S complex or the 26S proteasome of *Psmb9*^*G156D/G156D*^ mice, indicating genotype-dependent defect of immunoproteasome maturation (Fig. 2A). Catalytic activity of the 20S complex was decreased in a genotype-dependent manner (Fig. 2B). Meanwhile, in *Psmb9*^*G156D/+*^ and *Psmb9*^*G156D/G156D*^ mice, 26S proteasome activity was comparable to that of *Psmb9*^*+/+*^ mice and accumulation of ubiquitinated proteins was unremarkable (Fig. 2B,C). Thus, similar to the patients, not only *Psmb9*^*G156D/+*^ but also *Psmb9*^*G156D/G156D*^ mice showed severe defects mainly in 20S complex assembly and activity.

**Figure 2.**
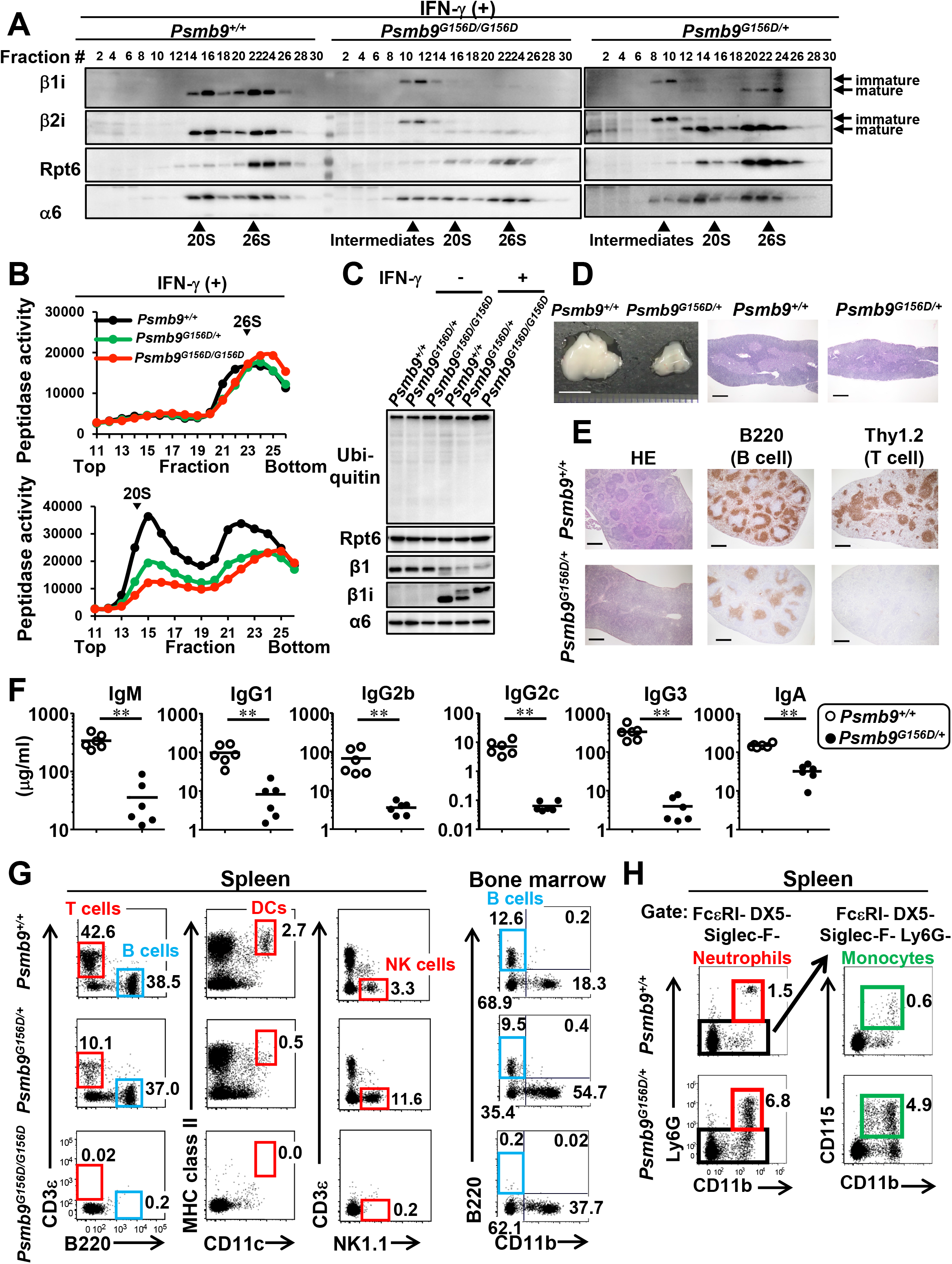
Analysis of mice carrying the PSMB9 G156D mutation. Panels A and B show the proteasome analysis of IFN-γ stimulated embryonic fibroblasts from the indicated mice. Cell extracts were prepared and fractionated by glycerol gradient centrifugation. Panel A shows immunoblot analysis of each fraction using Abs against the indicated proteins. Panel B shows chymotrypsin-like activity of each fraction, which was measured by using Suc-LLVY-AMC as a substrate in the absence (top) or presence (bottom) of 0.0025% SDS. Panel C shows the immunoblot analysis of embryonic fibroblast lysates using Abs against the indicated proteins. Panel D shows a photograph and histological images of the thymus. Scale bars in the leftmost and the other two panels represent 5 mm and 500 μm, respectively. Panel E shows cryosections of the spleen stained with hematoxylin and eosin (HE, left panel), anti-B220 Ab (middle panel), or anti-Thy1.2 Ab (right panel). Scale bars represent 500 μm. Panel F shows serum immunoglobulin levels. **p<0.01. Panel G shows FACS analysis of B and T cells, DCs, and NK cells, in the spleen or bone marrow. The numbers represent the percentages of the cells within the indicated gates or each quadrant among splenocytes or bone marrow cells. Panel H shows FACS analysis of neutrophils and monocytes in the spleen. Left and right panels show FcεRI-DX5-Siglec-F- and FcεRI-DX5-Siglec-F-Ly6G- cells, respectively. The numbers represent the percentages of neutrophils and monocytes among splenocytes.

We then performed immunological analysis. In *Psmb9*^*G156D/+*^ mice, thymus was small and the cortico-medullary junction was unclear (Fig. 2D). All thymocytes including their subsets were decreased in number (Fig. S4A). Histological analysis of the spleen also showed defective formation of follicles and decrease of B and T cells (Fig. 2E). Among splenocytes, B, as well as both CD4 and CD8 T cells, were decreased in number (Fig. S4B). In remaining CD8 T cells, central and effector memory T cells were increased, while naïve T cells were prominently decreased in percentages (Fig. S4C). Furthermore, serum levels of all Ig isotypes were severely decreased (Fig. 2F). Splenic dendritic cells (DCs) were also decreased in number and percentages (Figs. 2G, S4B). Meanwhile, NK cells were normal and neutrophils and monocytes were increased not only in spleen but also in BM (Figs. 2G, H and S4B). In *Psmb9*^*G156D/G156D*^ mice, thymus was hardly detected and spleen lacked T, B, NK and DCs (Fig. 2G). Thus, *Psmb9*^*G156D/+*^ mice showed combined immunodeficiency with increase of monocytes and neutrophils, and homozygosity of this mutation caused severer immunodeficient phenotype.

In mutant mice lacking proteasome subunits, generation of MHC class I-restricted Ag peptides is impaired, which leads to decreased expression of MHC class I and decrease of CD8 T cells.^16^ However, MHC class I expression was not impaired and processing and presentation of ovalbumin were also intact in splenic B cells of *Psmb9*^*G156D/+*^ mice, indicating that immunoproteasome of *Psmb9*^*G156D/+*^ mice functionally retained the MHC class I presentation activity (Fig. S5).

## DISCUSSION

We describe a novel *de novo* heterozygous missense mutation, p.G156D, in *PSMB9* coding an immunoproteasome subunit, β1i, in two unrelated patients that manifest characteristic autoinflammation, similar to so far described PRAAS-causing mutations, with immunodeficiency. Although the mutation led to defects in β1i maturation and formation and activity of immunoproteasome, it mainly caused formation and activity defects in the 20S complex and the 26S proteasome defects were mild enough at least to avoid ubiquitin accumulation, as shown by the analysis not only of the present two patients but also of the mice carrying the heterozygous or homozygous PSMB9 G156D mutation (Fig. 3). This is in contrast to published PRAAS cases, which mostly show impairment of both 20S and 26S formation and ubiquitin accumulation, accompanied with endoplasmic reticulum stress or type I interferonopathy.^3,8^ According to structural modeling analysis, PSMB9 G156 is located in the interface between two β rings and G156D mutation is considered to disturb interaction of two β rings, while keeping active site conformation. Meanwhile, most of published proteasome subunit mutations, as exemplified by T75M, A92T, A94P, K105Q, or G201V in PSMB8, are found at or near the active site and should affect the active site itself or its conformation (Fig. S6A), which is necessary for both 20S complex and 26S proteasome activities.^5^ Thus, PSMB9 G156D is a unique mutation in proteasome subunits which causes proteasome defects by its heterozygosity and affects the interaction of two β rings.

**Figure 3.**
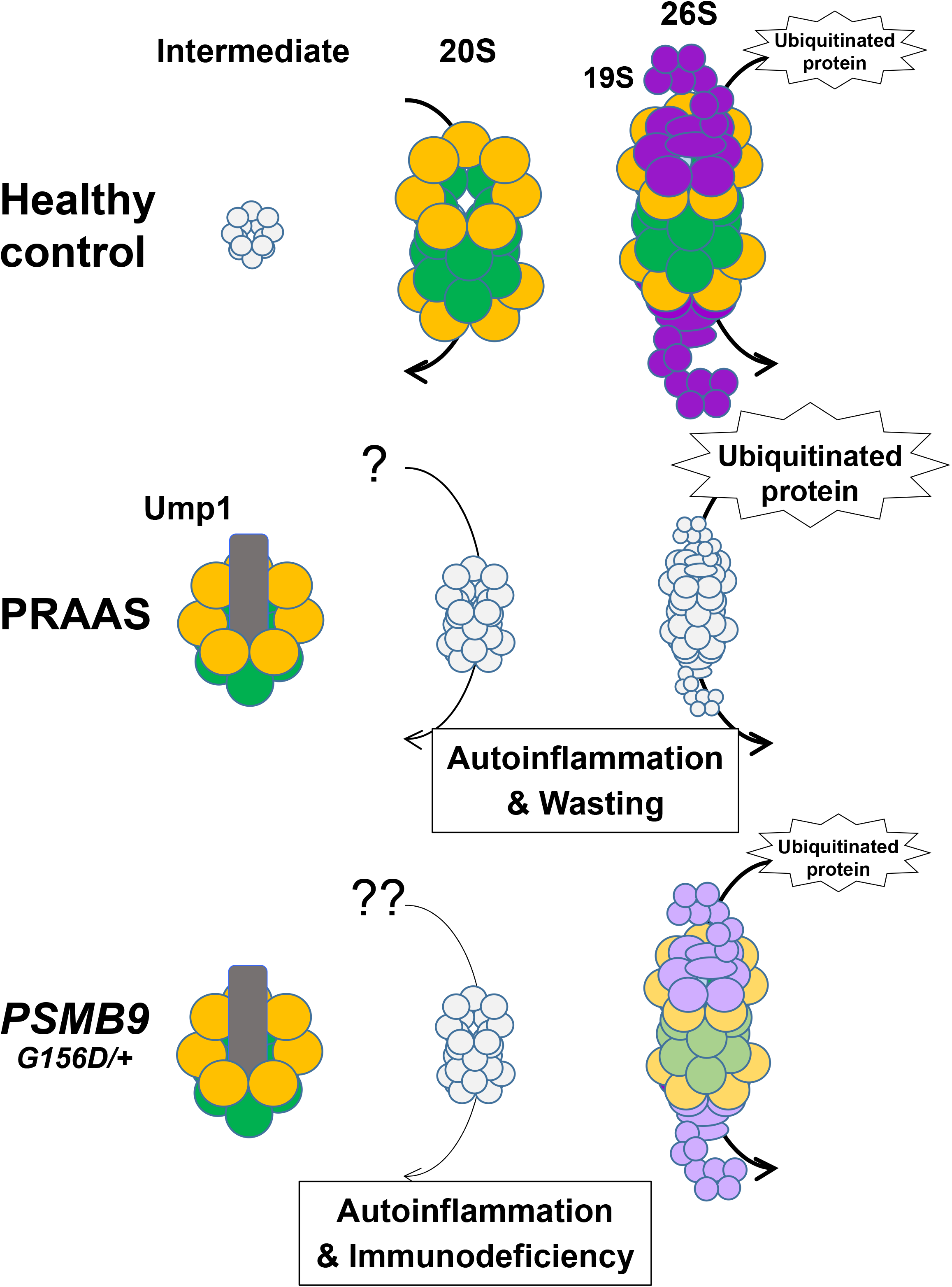
Schematic view of immunoproteasome assembly in PRAAS and patients. In healthy controls, the 20S complex of immunoproteasome is formed as αββα rings containing two α rings with α1-7 (orange circles) and two β rings with β1i, β2i, β3, β4, β5i, β6 and β7 (green circles). Then two 19S regulatory particles (purple circles) are incorporated to form the 26S proteasome, which is involved in degradation of ubiquitinated proteins. In PRAAS cases, the 20S and 26S immunoproteasome activities are impaired by defects of β ring assembly and/or of active site formation, which then lead to ubiquitin accumulation and autoinflammation with wasting. In patients carrying heterozygous PSMB9 G156D mutation, half immunoproteasomes and related intermediate forms were increased as in PRAAS cases. Notably, although the 20S complex formation was impaired, the 26S proteasome formation was less severely impaired and the 26S proteasome activity was kept at least to avoid apparent ubiquitin accumulation. Although the details are unclear, we can assume that this is caused by impaired interaction of β rings and that the 20S complex defects leads not only to autoinflammation but also to immunodeficiency, the latter of which is recapitulated in mice carrying the mutation. Quantity and activity of the 20S and 26S proteasome complexes are represented by size and color density of circles.

Immunodeficiency as well as proteasome defects, observed in two patients, were recapitulated in mice carrying the heterozygous PSMB9 G156D mutation. The mutant mice showed decrease of T, B cells and DCs, decrease of serum Ig levels and increase of neutrophils and monocytes. The phenotypes significantly overlap with that of patient 1, who showed decrease of B cells and serum IgG level and increase of monocytes (Fig. 2 and Table 1). Patient 2 also showed decrease of CD8 T in addition to γδ T and NK cells, although he was too young to be evaluated for B cell generation or serum IgG level. Despite the immunodeficiency, *Psmb9*^*G156D/+*^ mice appeared healthy without spontaneous development of autoinflammatory phenotypes. Some environmental factors, although not defined yet, might be necessary for *Psmb9*^*G156D/+*^ mice to manifest autoinflammation. Importantly, immunodeficient phenotypes in *Psmb9*^*G156D/G156D*^ mice were more prominent than those in *Psmb9*^*G156D/+*^ mice, suggesting the dosage effects of the PSMB9 G156D mutation. Although severe immunodeficiency should contribute to the early death of *Psmb9*^*G156D/G156D*^ mice, the direct cause for the death remains unknown.

So far, mutant mice lacking either or all of β1i, β2i, β5i, and β5t have been generated to show decrease of CD8 T cell numbers and responses.^16-21^ However, none of them exhibited combined immunodeficient phenotypes. Furthermore, CD8 T cell decrease observed in *Psmb9*^*G156D/+*^ mice is not caused by defective immunoproteasome activity to generate MHC class I-restricted antigens, as observed in the induced immunoproteasome subunit(s)-deficient mice,^16^ because MHC class I expression was not impaired and class I-restricted antigen presentation was normal in *Psmb9*^*G156D/+*^ mice (Fig. S5). It can be assumed that the mutation does not lead to mere decrease, but alteration or modification of immunoproteasome functions, which cannot be monitored by catalytic properties of conventionally used substrate polypeptides. Thus, although it remains unclear at present why PSMB9 G156D leads to immunodeficiency, mice with PSMB9 G156D mutation are the first and a unique model for analyzing proteasome defects.

Immunodeficient phenotype is also observed in mice carrying a missense mutation of β2i, which is introduced by N-ethyl-N-nitrosourea (ENU) mutagenesis.^22^ The mutation is described here as PSMB10 G209W, based on the amino acid number from the first methionine. *Psmb10*^*G209W*/+^ mice showed defects of both CD4 and CD8 T cells and *Psmb10*^*G209W/G209W*^ mice lacked B as well as T cells and manifested skin disorders with hyperkeratosis and infiltration of neutrophils. G209 is highly conserved among β2 and β2i across species (Fig. S6B) and G209W mutation led to decrease of the 20S complex formation while keeping the 26S proteasome formation, which is similar to the effects of PSMB9 G156D mutation.^22^ This speculation is supported by structural modeling analysis, showing that PSMB10 G209 is located at the interface of the two β rings and faced to β6 in the other half proteasome (Fig. S6A, C). Effects of PSMB10 G209W to induce immunodeficiency seem milder than those of PSMB9 G156D. It, however, can be reasonably assumed that the 20S complex formation is critical for immune cell homeostasis and that PSMB9 G156D and PSMB10 G209W should lead to immunodeficiencies through defective interaction of two β rings (Fig. 3). It remains unknown why the 26S proteasome formation and activity are relatively preserved, although 19S regulatory subunits might help facilitate the interaction.

Ump1 coded by *proteasome maturation protein (POMP)* is a chaperone used for proteasome assembly and heterozygous truncating mutations of this gene cause autoinflammatory phenotypes with type I IFN signature.^6^ The mutations also result in immune dysregulation, such as lymphocyte abnormalities showing CD4 T cell increase and B cell decrease and the authors propose a designation as POMP-related autoinflammation and immune dysregulation disease (PRAID). The manifestations are similar, but distinct from those of our cases (Table 1) and *Psmb9*^*G156D/+*^ mice, because the POMP mutations lead to both the 20S complex and the 26S proteasome defects with ubiquitin accumulation and hyper-γ-globulinemia with autoantibodies.^6^

Here we would propose a new designation “proteasome-associated autoinflammation and immunodeficiency disease (PRAID)” as an umbrella name for our cases as well as PRAAS and POMP-RAID, although contribution of autoinflammation, autoimmunity, and immunodeficiency is variable among mutations. PSMB9 G156D mutant mice are unique and useful to clarify not only the pathogenesis of proteasome dysfunctions but also novel homeostatic roles of immunoproteasome and to develop novel therapeutic maneuvers for PRAAS and immunodeficiency. It is also noteworthy that proteasome-related genes be candidate genes for screening patients with primary immunodeficiency of unknown etiology.

## Data Availability

Anyone who wishes to share, reuse, remix, or adapt this material must obtain permission from the corresponding authors.

## Acknowledgements

Supported by Health Labor Sciences Research Grants for Research on Intractable Diseases (grant 20316700 and 20317089 to H. Ohnishi) from the Ministry of Health, Labour and Welfare (MHLW) of Japan; by the Practical Research Project for Rare/Intractable Diseases under Grant Numbers JP15gk0110012 (to K. Yoshiura), JP16ek0109179 and JP19ek0109209 (to S. Okada), JP17ek0109100 (to N. Kanazawa), JP19ek0109199 (to, N. Kanazawa, H. Hemmi, N. Kinjo K. Yoshiura, and T. Kaisho), JP20ek0109480 (to H. Ohnishi and S. Okada), and Advanced Research and Development Programs for Medical Innovation (AMED-CREST) under Grant Number JP20gm1110003 (to S. Murata) from the Japan Agency for Medical Research and Development (AMED); by Grants-in-Aids for Scientific Research (A) (grant JP18H04022 to S. Murata), Scientific Research (B) (grant JP16H05355 and JP19H03620 to S. Okada, grant JP17H04088 and JP20H03505 to T. Kaisho), for Scientific Research (C) (grant JP15K09780 and JP19K08798 to N. Kanazawa, grant JP18K07840 to H. Ohnishi, grant JP19K07628 to I. Sasaki, grant JP18K07071 to H. Hemmi, grant JP16K10171 and JP19K08754 to K. Kunimoto), for Young Scientists (grant JP19K17293 to K. Izawa, grant JP20K16289 to T. Orimo, JP18K16096 to Y. Fukuda-Ohta), for Scientific Research on Innovative Areas (grant JP17H05799 and JP19H04813 to T. Kaisho, JP18H05500 to S. Murata), for Exploratory Research (grant JP17K19568 to T. Kaisho), for Young Scientists (B) (grant JP16K19585 to Y. Fukuda-Ohta), Research Activity start-up (grant JP19K23848 to T. Orimo), Promotion of Joint International Research from the Japan Society for the Promotion of Science (JP18KK0228 to S. Okada) from the Japan Society for the Promotion of Science; by the Uehara Memorial Foundation (to I. Sasaki and T. Kaisho); by Takeda Science Foundation (to H. Hemmi, T. Mizushima, I. Sasaki and T. Kaisho); by the Ichiro Kanehara Foundation for the promotion of Medical Sciences and Medical care (to H. Hemmi); by the Inamori Foundation (to I. Sasaki); by the Extramural Collaborative Research Grant of Cancer Research Institute, Kanazawa University; by a Cooperative Research Grant from the Institute for Enzyme Research, Joint Usage/Research Center, Tokushima University; by the Grant for Joint Research Program of the Institute for Genetic Medicine Hokkaido University; by the Grant for Joint Research Project of the Institute of Medical Science, the University of Tokyo; by the Program of the Network-type Joint Usage/Research Center for Radiation Disaster Medical Science; by Wakayama Medical University Special Grant-in-Aid for Research Projects.

Disclosure forms provided by the authors are available with the full text of this article.

We thank Dr. Masahiro Yamamoto for useful advice and discussion for the CRISPR/Cas9 method, Ms. Ikuko Hattori and Ms. Chihiro Nakai for technical assistance, and Ms. Aoi Tawaki-Matsumura and Ms. Akane Nishiwaki for secretarial assistance.

## SUPPLEMENTARY APPENDIX

### SUPPLEMENTARY METHODS

#### Approval for this study

This study was approved by the Ethics Committee of the University of the Ryukyus for Medical and Health Research Involving Human Subjects, the ethics boards of Gifu University, the Ethics Committee on Human Genome / Gene Analysis Research Nagasaki University, the Research Ethics Committee and the Animal Research Committee of Wakayama Medical University.

#### IFN score

Total RNA was extracted from human blood samples collected into PAXgene Blood RNA tubes (762165, Becton, Dickinson and Company) and was reverse transcribed by PrimeScript II 1^st^ strand cDNA synthesis kit (6210A, Takara). Quantitative real time PCR was performed with Taqman Gene Expression Master Mix (4369016, Applied Biosystems) and probes (*IFI27*, Hs01086370_m1; *IFIT1*, Hs00356631_g1; *RSAD2*, Hs01057264_m1; *SIGLEC1*, Hs00988063_m1; *ISG15*, Hs00192713_m1; *IFI44L*, Hs00199115_m1; *BACT*, Hs01060665_g1, Applied Biosystems) on StepOnePlus Real-Time PCR system (Applied Biosystems). The expression levels of each transcript were determined in triplicate and normalized to the level of β-actin. Results were shown relative to a single calibrator. Median fold change of expression levels of the six IFN-stimulated genes was used to calculate the IFN score for each patient. The score was regarded as positive if it exceeded +2SD of the average IFN score from healthy controls.

To calculate IFN score in mice, total RNA was extracted with RNeasy micro kit (QIAGEN) from *Psmb9*^*+/+*^ and *Psmb9*^*G156D/+*^ splenocytes and reverse transcribed by PrimeScript RT reagent Kit (RR037A, Takara). Quantitative real time PCR was performed with TB Green Premix Ex Taq II (RR820A, Takara) and specific primers for six IFN-stimulated genes on StepOnePlus Real-Time PCR system (Applied Biosystems). The primers were as follows: *Ifi27*: 5’-TTCCCCCATTGGAGCCAAG- 3’ and 5’- AGGCTGCAATTCCTGAGGC-3’, *Ifit1*: 5’-CTGAGATGTCACTTCACATGGAA-3’ and 5’- GTGCATCCCCAATGGGTTCT-3’, *Rsad2*: 5’-GCTTGTGAGATTCTGCAAGGA-3’ and 5’- GGCCAATCAGAGCATTAACCTG-3’, *Siglec1*: 5’- AGTGATAGCAACCGCTGGTTA-3’ and 5’- GCACAGGTAGGGTGTGGAAC-3’, *Isg15*: 5’- CTTTCTGACGCAGACTGTAGA-3’ and 5’- GGGGCTTTAGGCCATACTCC-3’, *Ifi44*: 5’-CCTGGTTCAGCAAACACGAGT-3’ and 5’- TGGCCTTGATGGAATATGTCCT-3’. The expression levels of each transcript were normalized to the levels of 18S ribosomal RNA, which were determined by TaqMan probes (TaqMan Gene Expression Assay, Applied Biosystems). Median fold change of the six IFN-stimulated genes was used to calculate the IFN score for *Psmb9*^*+/+*^ and *Psmb9*^*G156D/+*^ mice. The score was regarded as positive if it exceeded +2SD of the average IFN score from *Psmb9*^*+/+*^ mice.

#### Cytokine measurement by ELISA

Human serum samples were stored at −80°Cuntil assayed. The concentrations of TNF-α, IL- 1β, IL- 6, IL-18, IFN-α and IP-10 were measured with ELISA kits (Invitrogen for TNF-α, IL-1β, and IL-6, R&D for IP-10, MBL for IL-18, and PBL Assay Science for IFN-α).

#### Western blot analysis of human fibroblasts

SV40 transformed fibroblasts were stimulated with 1,000 IU/mL of IFN-γ for 15 min. The total cell lysates were subjected to SDS-PAGE. Expression of p-STAT1, STAT1 and β-actin was evaluated with anti-p-STAT1 (pY701) Ab (58D6, Cell Signaling), rabbit polyclonal anti-STAT1 Abs (SANTA CRUZ) and anti-β-actin Ab (AC-74, Sigma), respectively.

#### Immunohistochemical analysis of human samples

Six μm sections of biopsy samples from the patients’ skin lesion were stained with rabbit polyclonal anti-p-STAT1 Abs (GeneTex) and anti-ubiquitin Abs (DAKO).

#### Exome analysis

Exon fragments were enriched from genomic DNA samples of a patient and the parents using the SureSelect Human All Exon Kit V5 kit (Agilent Technologies), according to the manufacturer’s instructions. The prepared libraries were sequenced by the HiSeq2500 sequencer (Illumina) to obtain 100bp+100bp paired-end reads. The reads were mapped by the Novoalign software (Novocraft Technologies Sdn Bhd) on the hg19 human reference genome. The Genome Analysis Toolkit was used for following local realignment and variants call to obtain SNV and small insertion/deletion (indel) calls combined with in-house workflow management tool.^1,2^ Using data from the patient and the parents, called variants that can be de novo mutations in the patients were selected using an in-house tool. Selected variants were annotated by using the ANNOVAR software.^3^ Finally, the variants meet the following criteria were selected as “deleterious” mutations: 1) leads stop gain, stop loss, nonsynonymous mutation, or splice site mutation, 2) alternative allele frequencies in databases including Complete Genomics whole genome data, the 1000genome project, NHLBI GO ESP, the human genome variation database of the Japanese population^4^ and an in-house database) are all equal or less than 0.5%, 3) not included in Segmental duplication region defined in the UCSC genome browser.^5,6^

For target capture sequencing, ubiquitin-proteasome-system, autophagy and interferonopathy-related gene panel was purchased from Integrated DNA Technology (IDT) (Coralville, IA). Gene capture and library construction were performed using Hybridization capture of DNA libraries using xGen Lockdown Probes and Reagents (IDT) according to manufacture’s instructions. The libraries were sequenced by the MiSeq sequencer to obtain 300bp+300bp paired-end reads. For the reads, mapping, SNV/indel calling, annotation and narrowing were performed as with the exome analysis.

#### Predictive analysis of the protein structure

Structural models of wildtype and mutated immunoproteasomes were created based on the structure of the immunoproteasome [Protein Data Bank (PDB) ID code 3UNH] using the Swiss-Model server^7^ and CNS (Crystallography and NMR system) program.^8^

#### Mice

*Psmb9 G156D* mutant mice were generated by the CRISPR/Cas9 method. Briefly, guide RNA (gRNA) targeting to *Psmb9* exon5 (guide sequence; 5’- CTCCTACATTTATGGTTATG −3’) and mRNA for Cas9 endonuclease were generated by in vitro transcription using MEGAshortscript T7 (Life Technologies) and mMESSAGE mMACHINE T7 ULTRA kit (Life Technologies), respectively. The synthesized gRNA and Cas9 mRNA were purified with MEGAclear kit (Life Technologies). A single-stranded oligodeoxynucleotides (ssODN) containing the c.G467A mutation (5’- CATCTGTGGTGAAACGCCGGCACTCCTCAGGGGTCATGCCTGGCTTATAAGCTGCGTCCACATAATCATAAATGTAGGAGCTTCCGGAACCGCCGATGGTAAAGGGCTGTCGAATTAGCATCCCTCCCATG −3’) was synthesized by Integrated DNA Technologies. To generate mutant mice, female B6C3F1 mice were superovulated and mated with C57BL/6N males (Clea Japan). Fertilized one-cell-stage embryos were injected with gRNA, Cas9 mRNA, and ssODN. The c.G467A mutation was confirmed by sanger sequencing, and the mutant mice were further backcrossed to C57BL/6N mice for more than 6 generations. Eight to sixteen weeks old mutant mice and their littermates were analyzed.

In order to generate OVA-expressing *Psmb9 G156D* mice, *Psmb9 G156D* mutant mice were crossed with membrane-bound OVA expressing transgenic mice (C57BL/6-Tg(CAG-OVA)916Jen/J, the Jackson laboratory)^9^.

#### Cell preparation

Immortalized B cells were generated by infecting patient 1-derived peripheral blood cells with Epstein-Barr virus. Fibroblasts were generated from patient 2-derived skin sample. The control human fibroblasts were purchased from the Japanese Collection of Research Bioresources, Osaka, Japan: SF- TY; TOYOBO, Osaka, Japan:106-05a; and KURABO, Osaka, Japan: KF-4109. The fibroblasts were transformed with origin-defective mutant of SV40 virus carrying wild-type T antigen.

For preparing mouse embryonic fibroblasts, E13.5-14.5 embryos were dissected, cut into small pieces, and soaked in 0.25% trypsin/EDTA (Nacalai Tesque) at 37°C with shaking for 30 min. The cells were then suspended by pipetting, filtrated with 100μm pore nylon mesh, and plated on a 15cm dish per embryo, and cultured in DMEM supplemented with 10% FBS. Cells cultured during days 5- 10 were used for experiments.

#### Flow cytometry

For the analysis of human lymphocyte subsets, peripheral blood samples were stained with fluorochrome-conjugated or biotinylated Abs for human CD3 (UCHT1), CD4 (13B8.2), CD8 (B9.11), CD16 (3G8), CD19 (J3-119), CD20 (B9E9), CD25 (B1.49.9), CD27 (1A4CD27), CD45RA (ALB11), CD45RO (UCHL1), CD56 (N901), CD127 (R34.34), TCRαβ (IP26A), TCRγδ (IMMU510) (IOTest; Beckman Coulter), CD3(HIT3a) and IgD (IA6-2) (BD Biosciences). Cells were analyzed on a Navios EX flow cytometer (Beckman Coulter).

Single cell suspensions of splenocytes and bone marrow cells from *Psmb9*^*+/+*^, *Psmb9*^*G156D/+*^, *and Psmb9*^*G156D/G156D*^ were incubated with an Ab against CD16/32 (2.4G2, TONBO) to block non-specific binding of Abs to Fc receptors. Then, the cells were stained with fluorochrome-conjugated or biotinylated Abs for murine CD3ε (145-2C11) (eBioscience), CD11b (M1/70), H-2D^b^ (KH95), Ly6C (HK1.4), Ly6G (1A8) (BioLegend), B220 (RA3-6B2), CD8α (53-6.7), CD44 (IM7), I-A/I-E (MHC class II, M5/114.15.2) (TONBO), CD62L (MEL-14), NK1.1 (PK136) (BD Bioscience), CD4 (GK1.5 or RM4-5) (BD Bioscience and BioLegend), CD11c (N418 or HL3) (BD Bioscience and TONBO), H-2K^b^ (AF6-88.5.5.3) (eBioscience and BD Bioscience). Biotinylated Abs were visualized by fluorochrome-conjugated streptavidin (BD Bioscience, BioLegend and eBioscience). Dead cells were excluded by staining with Fixable Viability Dye (eBioscience). To detect MHC class I-mediated presentation of endogenous OVA, splenocytes were stained with an Ab (25-D1.16) (eBioscience) against OVA-derived peptide/H2-K^b^ complex. Cells were analyzed on a FACS Verse or FACS Aria II (BD Biosciences) and data analyzed with FlowJo software (BD Bioscience).

#### Cell lysates, Glycerol gradient analysis, measurement of proteasomal activity, and western blotting

Cells were lysed in buffer containing 25 mM Tris-HCl (pH7.5), 0.2% NP-40, 1 mM DTT, 2 mM ATP, 5 mM MgCl_2_, and 1 mM phenylmethylsulfonyl fluoride (PMSF) and clarified by centrifugation at 20,000 x g for 10min at 4°C. For glycerol gradient centrifugation analysis, clarified cell lysates were subjected to 8-32% (v/v) glycerol linear density gradient centrifugation (22 h, 83,000 x g) and separated into 32 fractions, followed by measurement of peptidase activity of each fraction as described previously.^10^ Subsequently, peptidase activity was measured using a fluorescent peptide substrate, succinyl-Leu-Leu-VI-Tyr-7-amido-4-methylcoumarin (Suc-LLVY-AMC), for chymotrypsin-like activity as previously reported.^10^ The Abs against Ump1, β1i, β2i, β5i, β1, β2, β5, α2, α6 and polyubiquitin were described previously.^10-13^

#### Histology in mice

Thymus and spleen were fixed with 4% paraformaldehyde and embedded in OCT compound (Sakura Finetek) or FSC22 frozen section compound (Leica Microsystems), and 5 μm cryosections were prepared. The sections were stained with hematoxylin and eosin (Muto Pure Chemicals). To detect B and T cells, sections were stained with biotinylated Abs against B220 (RA3-6B2) or Thy1.2 (30-H12) after blockade of endogenous peroxidase with 3% H_2_O_2_/methanol. Biotinylated Abs were developed with VECTASTAIN ABC Standard kit and ImmPACT DAB (Vector Laboratories). Hematoxylin was used as counterstain.

#### Measurement of serum Ig levels in mice

Serum Ig levels were measured by ELISA developed in hand. In brief, serum samples were incubated with ELISA plate coated with goat anti-mouse IgM, IgG1, IgG2b, IgG2c, IgG3, or IgA Abs (Southern Biotech) and detected with biotinylated goat Abs against each class (Southern Biotech) and streptavidin-conjugated alkaline phosphatase. The plate was developed by alkaline phosphatase buffer (50mM NaHCO_3_, 10mM MgCl_2_, pH9.8) containing phosphatase substrate tablet (S0942, Sigma-Aldrich). Purified mouse IgM, IgG1, IgG2b, IgG2c, IgG3 (Southern Biotech) were used as standard.

**Figure S1.**
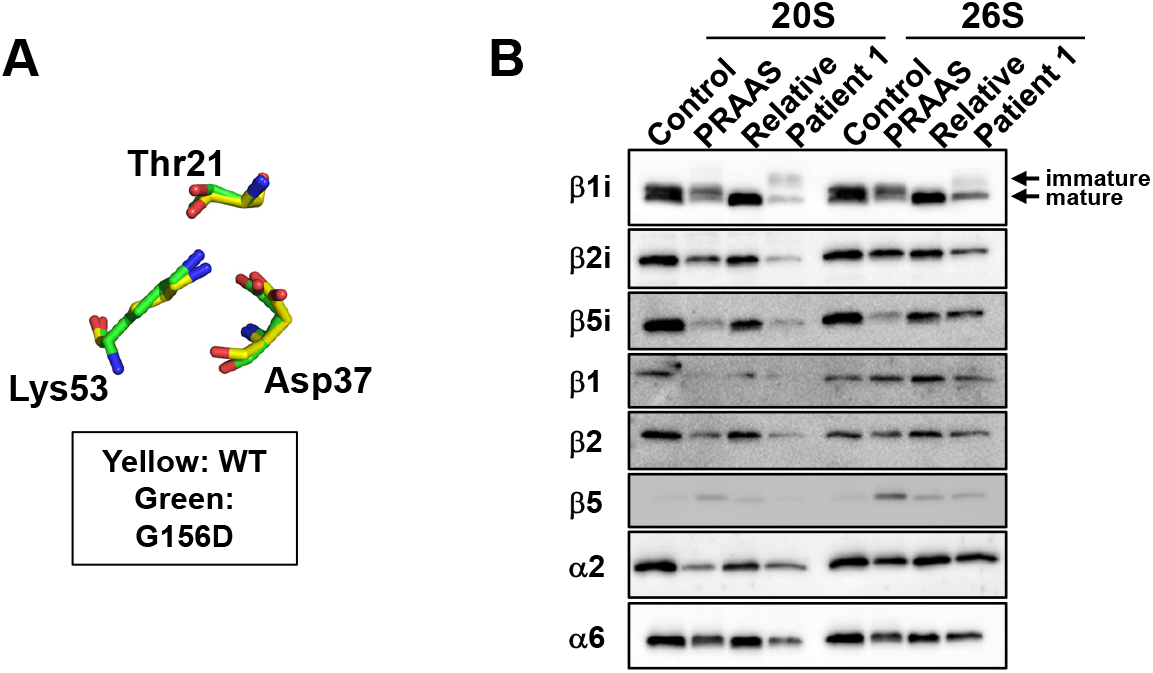
Effects of PSMB9 G156D on the activity and composition of proteasome. Panel A shows the catalytic residues of wildtype PSMB9 and PSMB9 G156D. Thr21 is the most important catalytic residue within the N terminus of mature β1i. Nitrogen and oxygen atoms are shown in blue and red, respectively. The wildtype and G156D structure is shown in yellow and green, respectively. Panel B shows immunoblot analysis of the 20S complex and the 26S proteasome fractions of a healthy control, a PRAAS patient, patient 1 and patient 1’s relative using Abs against the indicated proteins.

**Figure S2.**
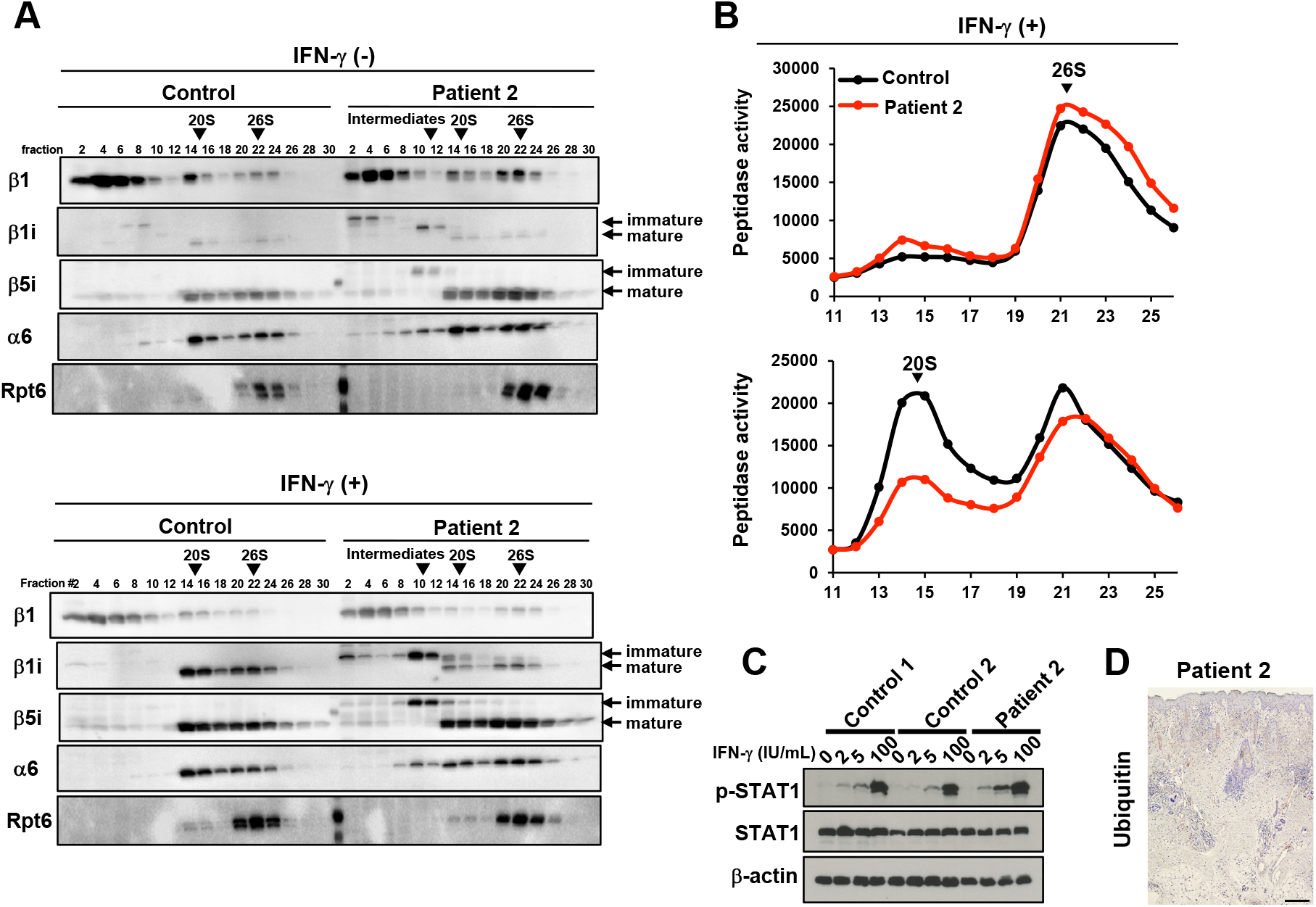
Composition and activity of proteasome in patient 2. Panels A and B show the proteasome analysis of unstimulated or IFN-γ-stimulated skin fibroblasts from a healthy control and patient 2. Cell extracts were prepared and fractionated by glycerol gradient centrifugation. Panel A shows immunoblot analysis of each fraction using Abs against the indicated proteins. Panel B shows chymotrypsin-like activity of each fraction, which was measured by using Suc-LLVY-AMC as a substrate in the absence (top) or presence (bottom) of 0.0025% SDS. Panel C shows the STAT1 phosphorylation assay with SV40-transformed dermal fibroblasts of control subjects and Patient 2. The expression levels of IFN-γ induced p-STAT1 were enhanced in patient 2 compared to control subjects. Panel D shows the skin biopsy sample of patient 2 stained with anti-ubiquitin Ab. A scale bar represents 100 μm.

**Figure S3.**
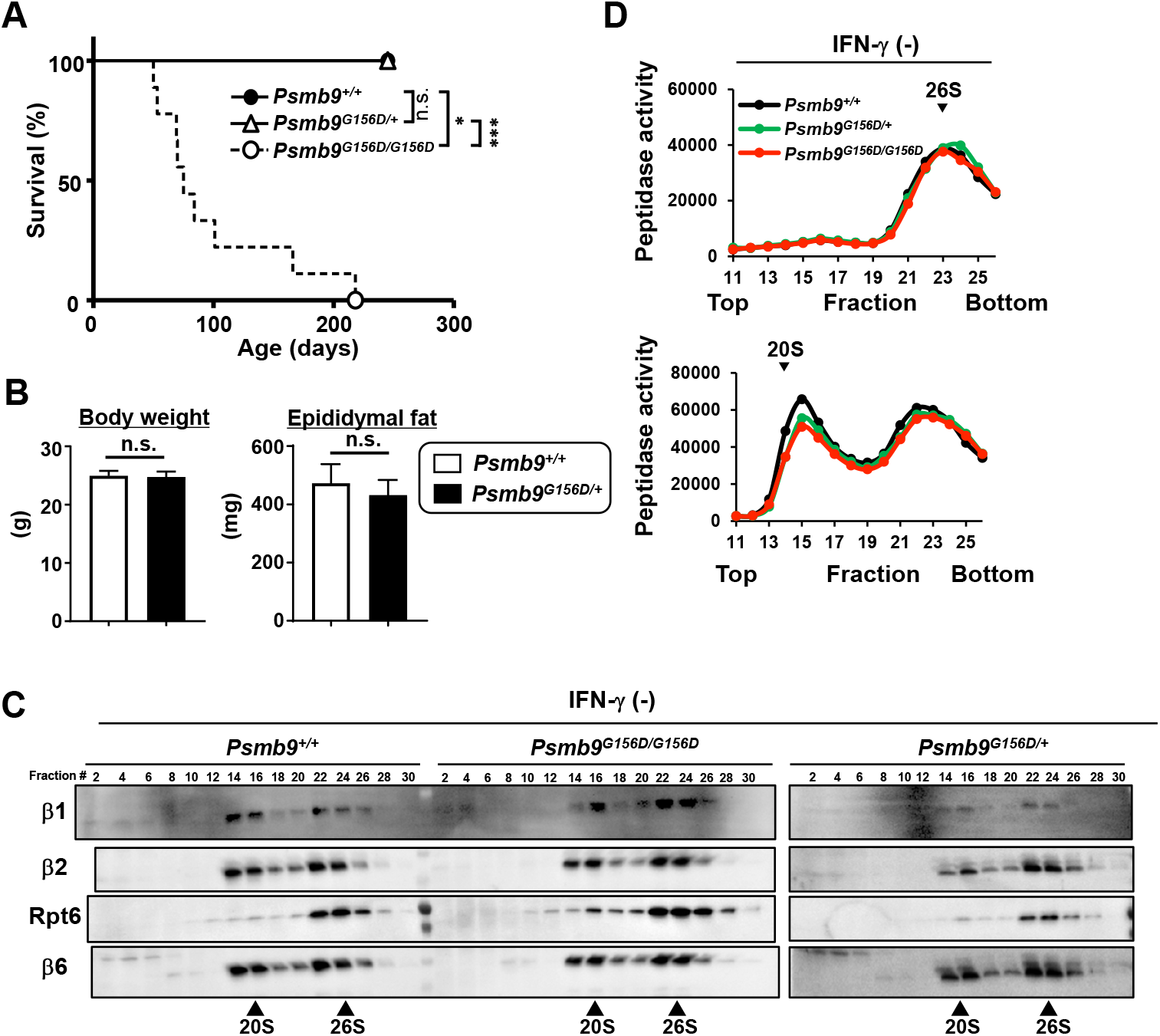
General status and proteasome activity in mice carrying the PSMB9 G156D mutation. Panel A shows cumulative survival curves (*Psmb9*^*+/+*^, n=2; *Psmb9*^*G156D/+*,^ n=6; *Psmb9*^*G156D/G156D*^, n=9). *p<0.05; ***p<0.001; n.s., not significant. Panel B shows body weight (left panel) and epididymal fat weight (right panel) (*Psmb9*^*+/+*^, n=4; *Psmb9*^*G156D/+*^, n=7). n.s., not significant. Panels C and D show the proteasome analysis of unstimulated embryonic fibroblasts from the indicated mice. Cell extracts were prepared and fractionated by glycerol gradient centrifugation. Panel C shows immunoblot analysis of each fraction using Abs against the indicated proteins. Panel D shows chymotrypsin-like activity of each fraction, which was measured by using Suc-LLVY-AMC as a substrate in the absence (top) or presence (bottom) of 0.0025% SDS.

**Figure S4.**
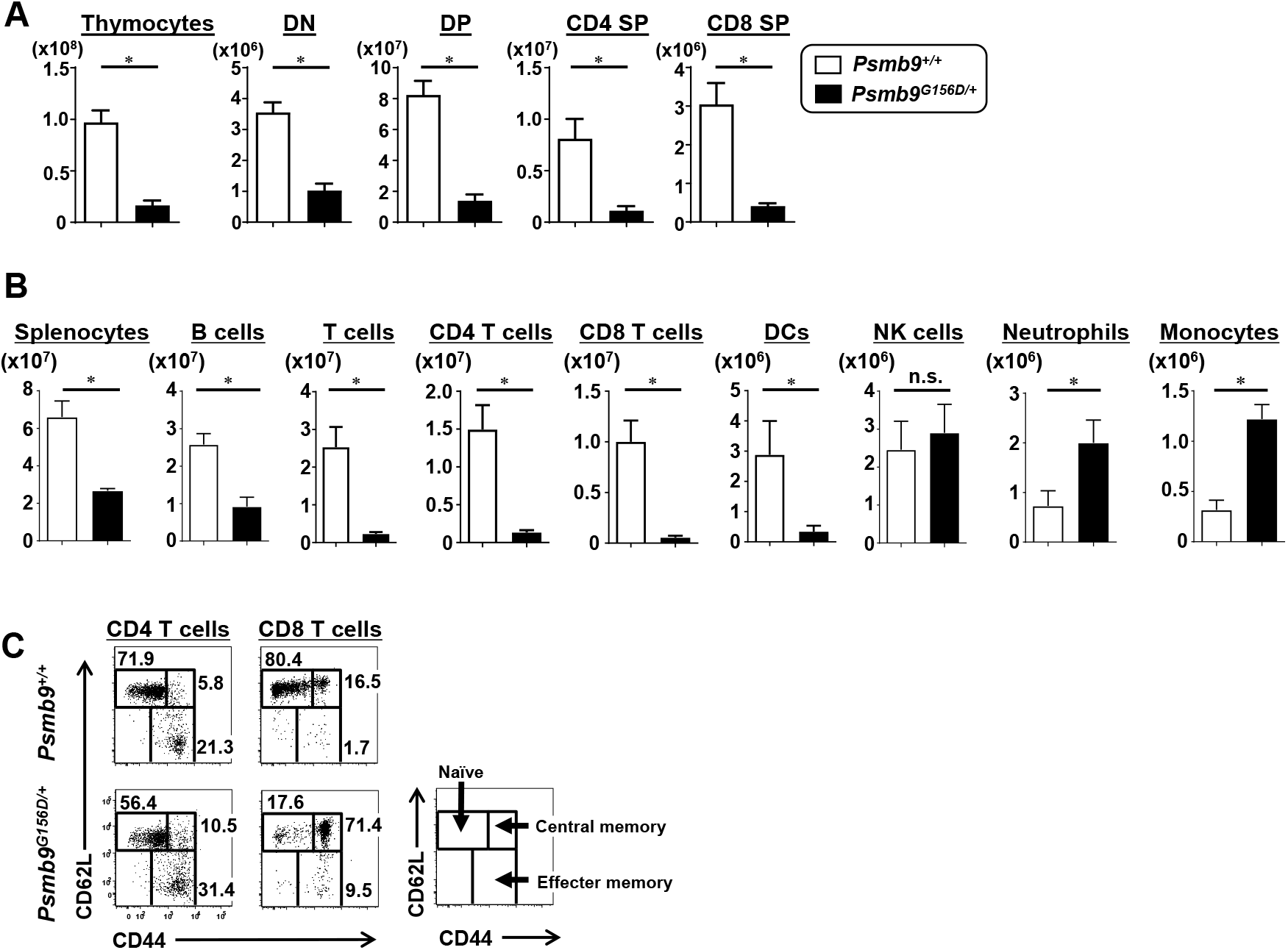
Thymus and spleen cellularity in mice carrying the PSMB9 G156D mutation. Panel A shows numbers of total thymocytes, CD4 CD8 double negative (DN), CD4 CD8 double positive (DP), CD4 single positive (CD4 SP), and CD8 single positive (CD8 SP) cells (*Psmb9*^*+/+*^, n=4; *Psmb9*^*G156D/+*^, n=4). *p<0.05. Panel B shows numbers of total splenocytes, B220+ B cells, CD3ε+ T cells, MHC class II+CD11c+ DCs, CD3e-NK1.1+ NK cells, FcεRI-DX5-Siglec-F-Ly6G+CD11b+ neutrophils, and FcεRI- DX5-Siglec-F-Ly6G-CD115+CD11b+ monocytes (*Psmb9*^*+/+*^, n=4; *Psmb9*^*G156D/+*^, n=4). *p<0.05. n.s., not significant. Panel C shows FACS analysis of CD44 and CD62L expression on splenic CD3ε+ T cells. The numbers in the dot plots indicate the percentages of the cells within the indicated gates. Representative data were shown.

**Figure S5.**
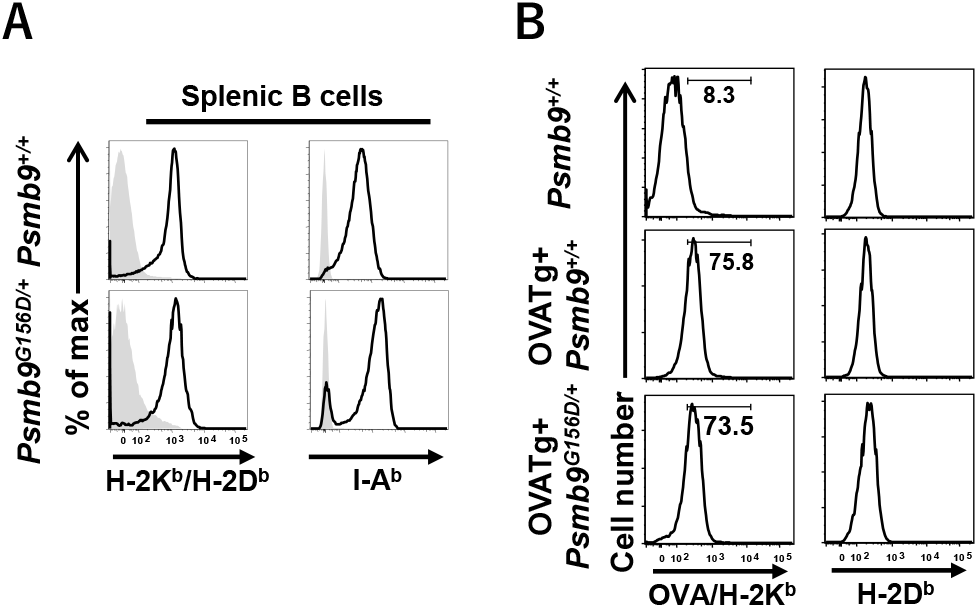
MHC class I antigen presentation in mice carrying the PSMB9 G156D mutation. Panel A shows representative histogram of MHC class I (H-2K^b^/H-2D^b^) and MHC class II (I-A^b^) expression on splenic B cells. Filled and open histograms indicate data stained with isotype matched Abs and specific Abs, respectively. Panel B shows MHC class I-associated presentation of endogenous antigen. Expression of an OVA-derived OVA_257-264_ peptide, SIINFEKL, on H-2K^b^ (OVA/H-2K^b^) and H-2D^b^ was shown.

**Figure S6.**
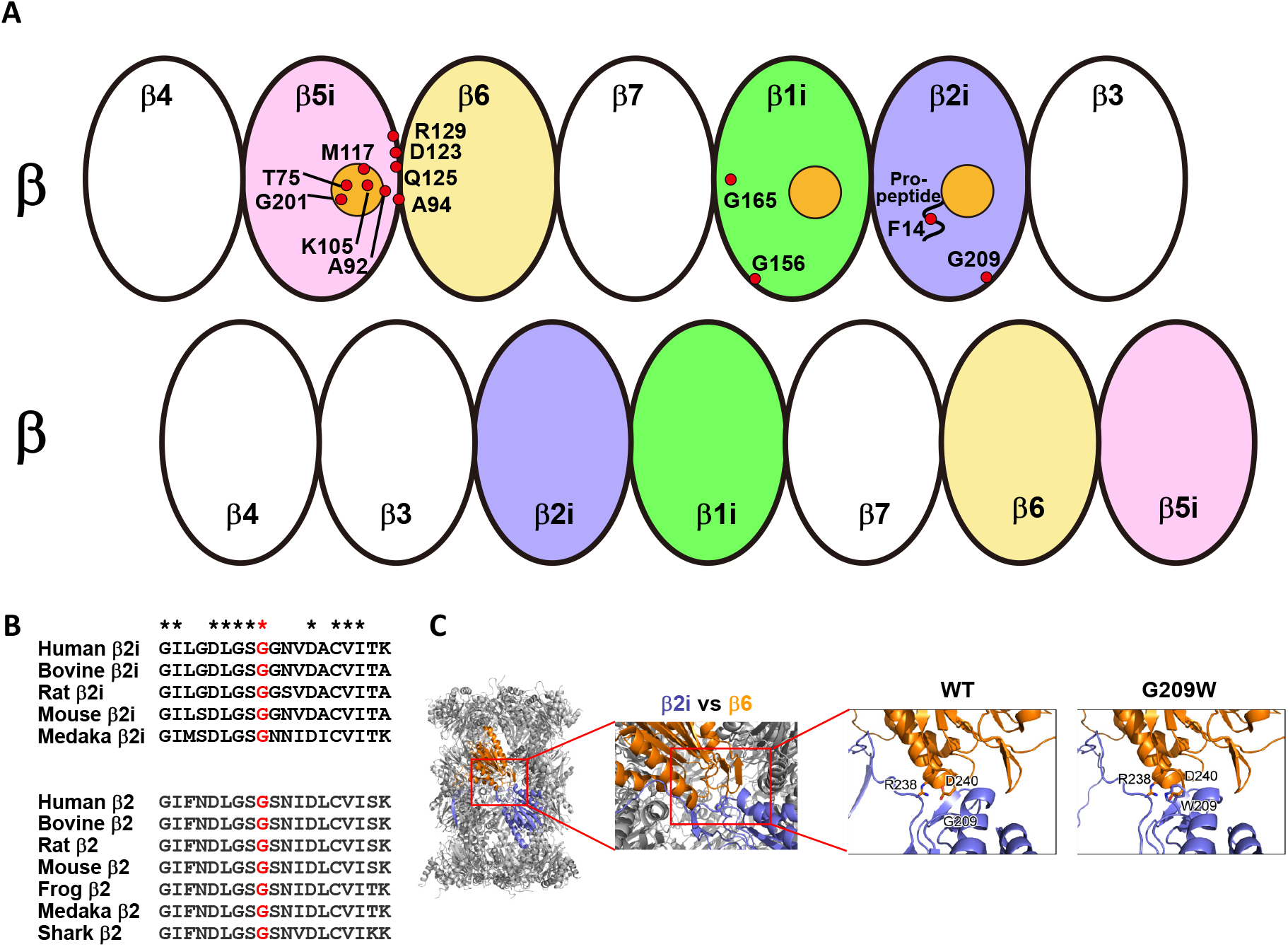
Characterization of proteasome subunit missense mutations. Panel A shows locational distribution of published missense mutation sites on the schematic model of β rings in the immunoproteasome (https://infevers.umai-montpellier.fr/web/index.php). The mutant residues are represented by red circles. Catalytic active sites in β1i, β2i and β5i are marked with orange circles. Notably, PSMB9 G156 and PSMB10 G209 are at the interface of two β rings, while most of mutated residues in PSMB8 are at or around active sites. Panel B shows multiple alignment of PSMB10 (β2i) and PSMB2 (β2) in various species and high conservation of G209 in murine PSMB10 (red). Panel C shows the structure model of wildtype and PSMB10 G209W mutant of the 20S complex, based on the β2i-subunit structure [Protein Data Bank (PDB) ID code 3UNH]. Overall structure of the 20S complex is shown as a ribbon model. β2i and β6 subunits are shown as light blue and orange, respectively. W209 and some of the potential interacting residues of PSMB10 G209W mutant are shown in stick representation.

**Table S1.** Immunophenotype of the studied individuals.

|  | Patient 1<br>(6-10 years old) | Relative of patient 1<br>(11-15 years old) | references<br>(1-15 years old) | Patient 2<br>(3-5 months old) | references<br>(0-1 year old) |
| --- | --- | --- | --- | --- | --- |
| WBC (/μl) | **15200 ▲** | 5000 | 4500-13500 | **3830 ▼** | 6000-17500 |
| Lymphocytes (/μl) | **988 ▼** | 2485 | 1500-6500 | **1226 ▼** | 4000-13500 |
| Neutrophils (/μl) | **13072 ▲** | 2035 | 1800-8000 | 2490 | 1000-8500 |
| Monocytes (/μl) | **836 ▲** | 220 | 180-540 | **77 ▼** | 300-875 |
| Lymphocyte Subsets |  |  |  |  |  |
| CD3+ | **80.2% ▲** | 65.8% | 65.7±6.6% | 70.5% | 69.8±14.1% |
| CD4+ | 48.4% | 40.7% | 42.6±7.6% | 60.1% | 49.1±16.4% |
| CD8+ | **29.2% ▲** | 23.9% | 22.1±6.9% | **7.8% ▼** | 17.8±7.5% |
| TCRαβ+ | **74.0% ▲** | 65.8% | 57.3±10.5% | 73.8% | 63.9±12.8% |
| TCRγδ+ | **1.2% ▼** | 4.9% | 5.7%±3.0% | **0.2% ▼** | 3.4±1.1% |
| CD4+, CD45RA+ | 25.7% | 28.6% | 32.7±7.9% | 57.6% | 45.0±16.6% |
| CD4+, CD45RO+ | **25.5% ▲** | 14.6% | 11.4±3.5% | **3.5% ▼** | 6.3±2.6% |
| CD8+, CD45RA+ | **27.4% ▲** | 19.0% | 19.7±5.6% | 7.9% | 15.9±6.5% |
| CD8+, CD45RO+ | 4.3% | 6.2% | 3.8±2.7% | 0.4% | 2.5±2.6% |
| CD25+, CD127dim (in CD4+) | 7.9% | 9.0% | 9.1±2.2% | 8.3% | 8.2±2.3% |
| CD3-, CD56+, CD16+ | 7.1% | 7.5% | 9.6±4.3% | **0.3% ▼** | 10.2±7.5% |
| CD19+ | **1.0% ▼** | 14.4% | 11.9±4.9% | 12.8% | 10.9±5.3% |
| CD20+ | **0.9% ▼** | 14.3% | 12.2±4.9% | 12.9% | 10.2±4.9% |
| CD27+, IgD- (in CD20+) | **47.4% ▲** | 7.9% | 7.5±4.5% | 0.7% | 1.8±1.5% |
| CD27+, IgD+ (in CD20+) | **26.3% ▲** | 11.1% | 11.2±6.8% | **11.5% ▲** | 7.0±2.2% |
| CD27-, IgD+ (in CD20+) | **10.5% ▼** | 77.3% | 76.5±10.8% | 87.1% | 87.7±3.3% |

Bold numbers indicate abnormal range.

**Table S2.**
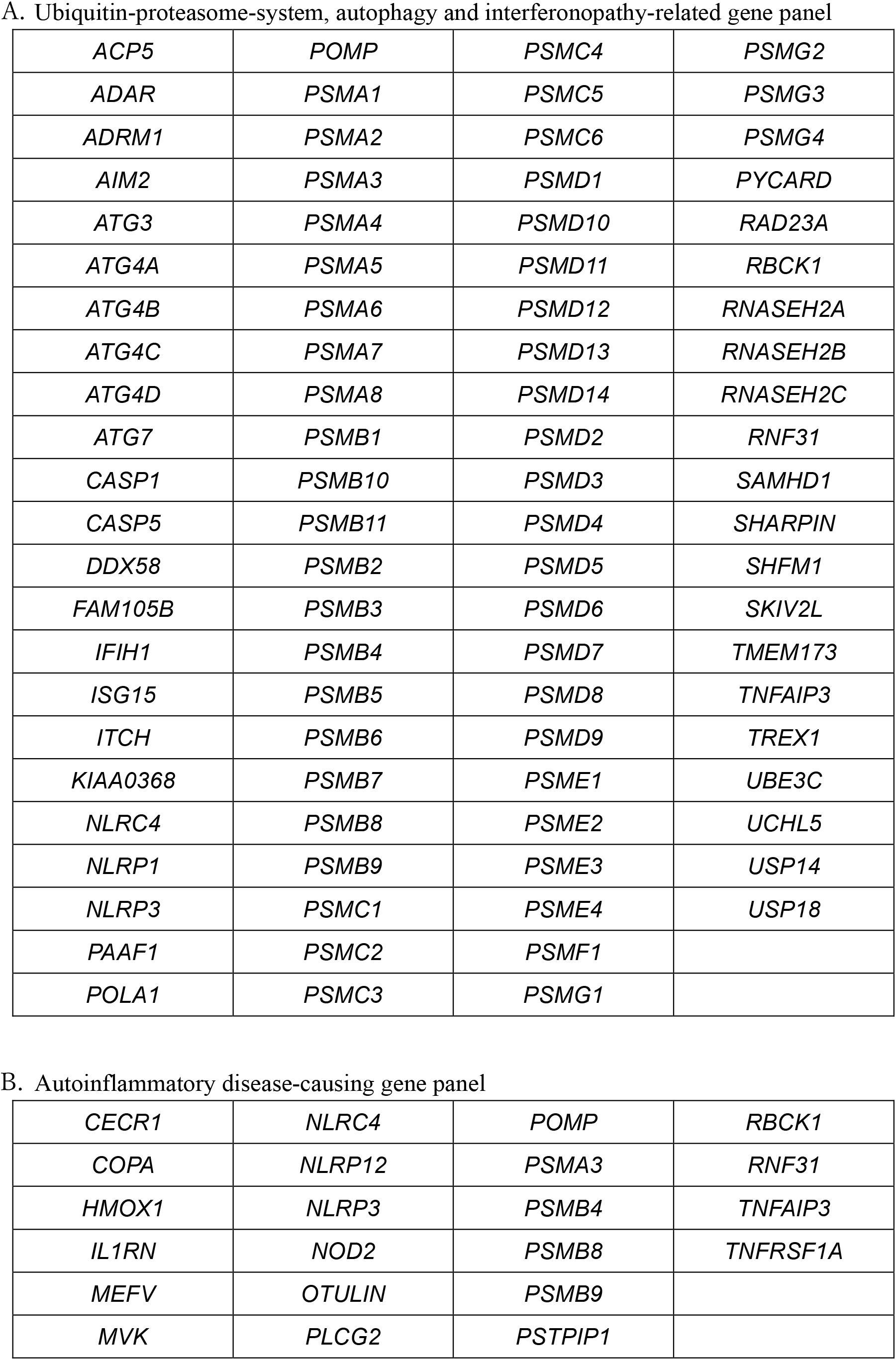
Target gene panels for sequencing.

**A. Ubiquitin-proteasome-system, autophagy and interferonopathy-related gene panel**
|  |  |  |  |
| --- | --- | --- | --- |
| *ACP5* | *POMP* | *PSMC4* | *PSMG2* |
| *ADAR* | *PSMA1* | *PSMC5* | *PSMG3* |
| *ADRM1* | *PSMA2* | *PSMC6* | *PSMG4* |
| *AIM2* | *PSMA3* | *PSMD1* | *PYCARD* |
| *ATG3* | *PSMA4* | *PSMD10* | *RAD23A* |
| *ATG4A* | *PSMA5* | *PSMD11* | *RBCK1* |
| *ATG4B* | *PSMA6* | *PSMD12* | *RNASEH2A* |
| *ATG4C* | *PSMA7* | *PSMD13* | *RNASEH2B* |
| *ATG4D* | *PSMA8* | *PSMD14* | *RNASEH2C* |
| *ATG7* | *PSMB1* | *PSMD2* | *RNF31* |
| *CASP1* | *PSMB10* | *PSMD3* | *SAMHD1* |
| *CASP5* | *PSMB11* | *PSMD4* | *SHARPIN* |
| *DDX58* | *PSMB2* | *PSMD5* | *SHFM1* |
| *FAM105B* | *PSMB3* | *PSMD6* | *SKIV2L* |
| *IFIH1* | *PSMB4* | *PSMD7* | *TMEM173* |
| *ISG15* | *PSMB5* | *PSMD8* | *TNFAIP3* |
| *ITCH* | *PSMB6* | *PSMD9* | *TREX1* |
| *KIAA0368* | *PSMB7* | *PSME1* | *UBE3C* |
| *NLRC4* | *PSMB8* | *PSME2* | *UCHL5* |
| *NLRP1* | *PSMB9* | *PSME3* | *USP14* |
| *NLRP3* | *PSMC1* | *PSME4* | *USP18* |
| *PAAF1* | *PSMC2* | *PSMF1* |  |
| *POLA1* | *PSMC3* | *PSMG1* |  |
| **B. Autoinflammatory disease-causing gene panel** |  |  |  |
| *CECR1* | *NLRC4* | *POMP* | *RBCK1* |
| *COPA* | *NLRP12* | *PSMA3* | *RNF31* |
| *HMOX1* | *NLRP3* | *PSMB4* | *TNFAIP3* |
| *IL1RN* | *NOD2* | *PSMB8* | *TNFRSF1A* |
| *MEFV* | *OTULIN* | *PSMB9* |  |
| *MVK* | *PLCG2* | *PSTPIP1* |  |

**Table S3.**
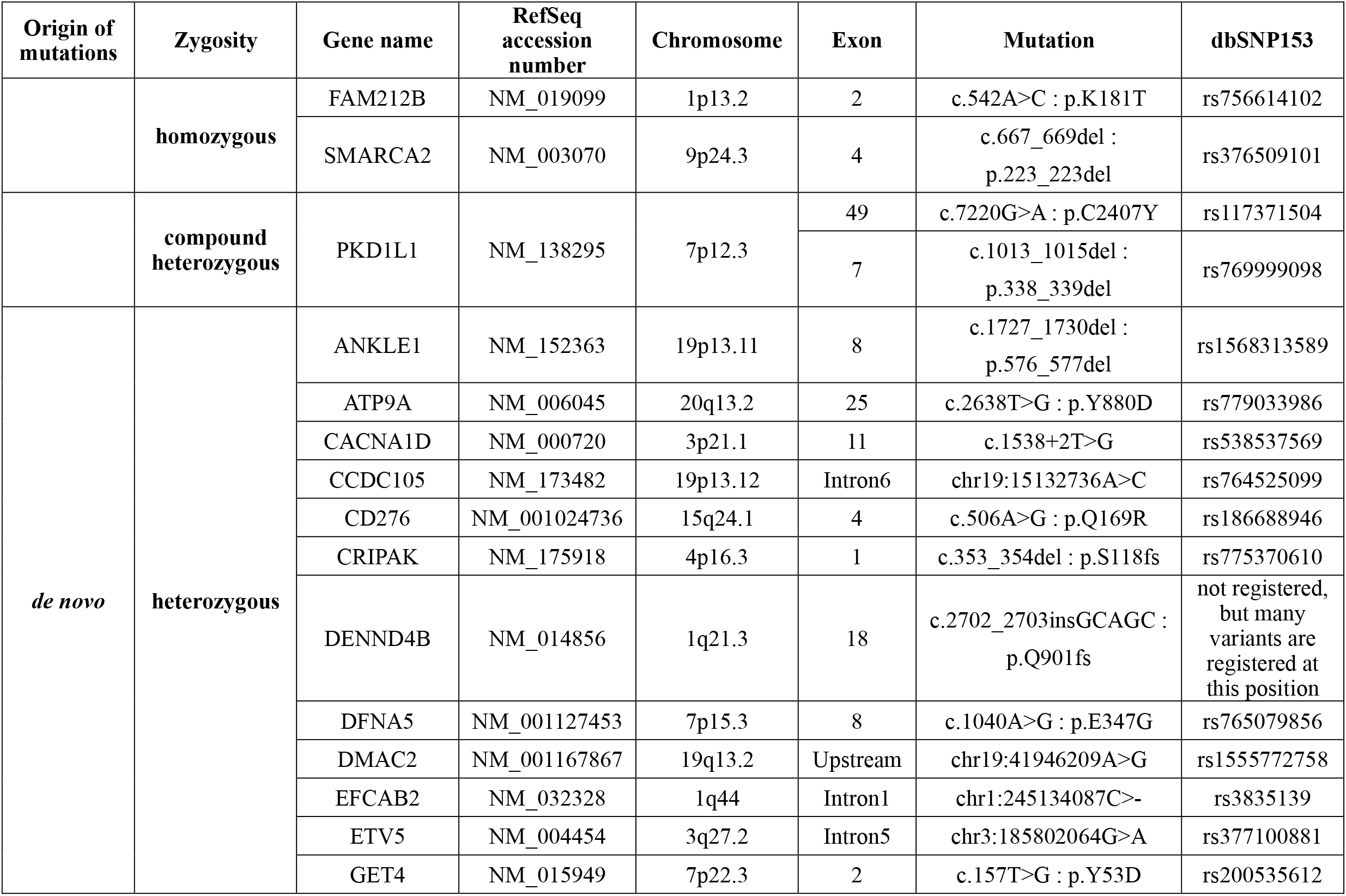
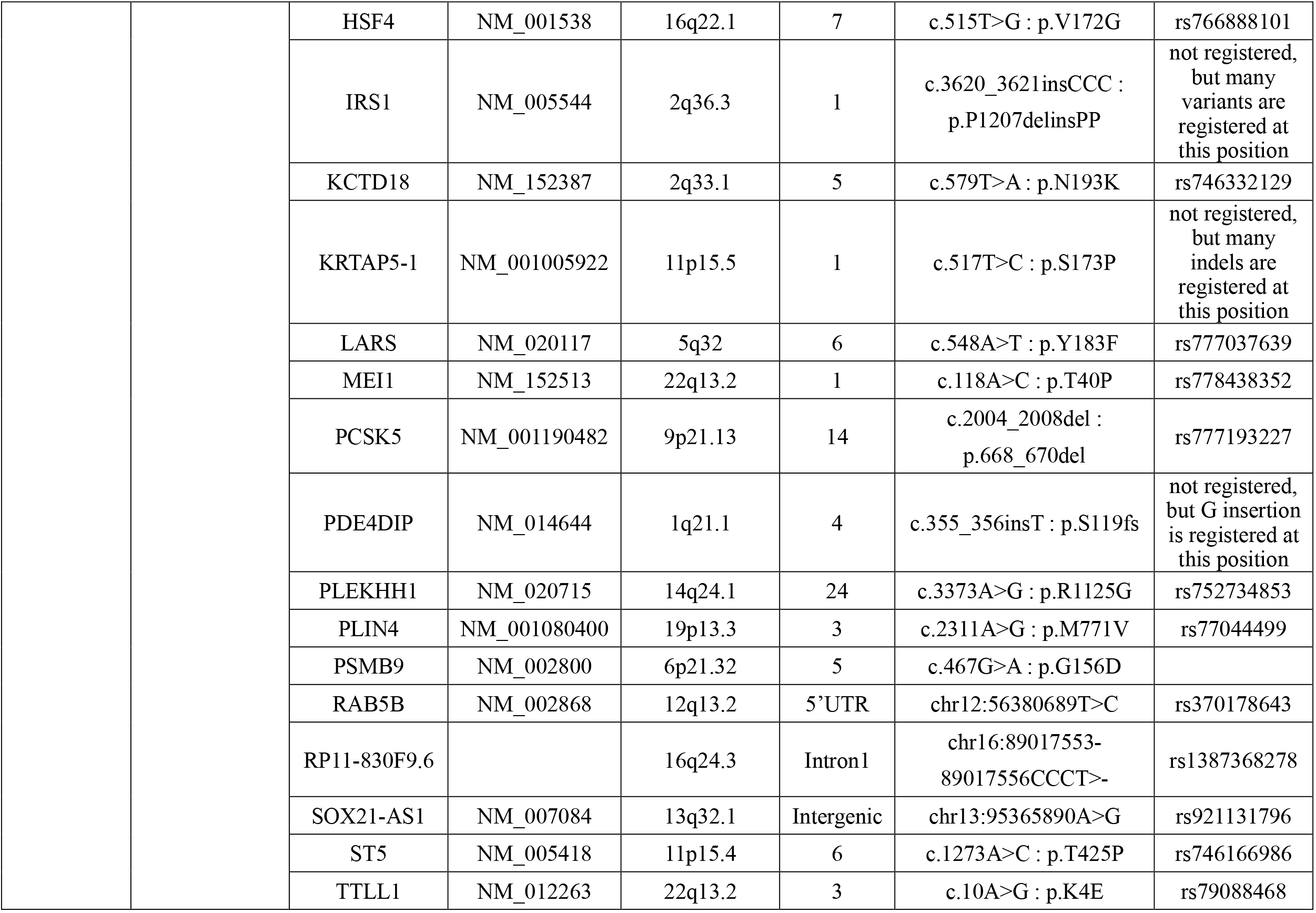
Candidate gene mutations identified by the exome sequencing of patient 1 and the parents.

**Table S4.** Candidate gene mutations identified by the exome sequencing of patient 2 and the parents.

| Origin of mutations | Zygosity | Gene name | RefSeq accession number | Chromosome | Exon | Mutation | dbSNP153 |
| --- | --- | --- | --- | --- | --- | --- | --- |
|  | **homozygous** | ALMS1 | NM_015120 | 2p13.1 | 1 | c.41_42insGGA :<br>p.E14insE | rs55889738 |
|  |  | TBP | NM_003194 | 6q27 | 3 | c.231delG : p.Q77fs | rs770128377 |
|  | **compound heterozygous** | IGHMBP2 | NM_002180 | 11q13.3 | 3<br>10 | c.344C>T : p.T115M<br>c.1483G>A : p.G495S | rs181657861<br>rs201964221 |
| *de novo* | **heterozygous** | AC004980.7 |  | 7q11.23 | Intron2 | chr7:76247636G>T | rs367687702 |
|  |  | BTN3A3 | NM_006994 | 6p22.2 | 4 | c.197G>A : p.R66K | rs1266759094 |
|  |  | CEP20 | NM_001304499 | 16p13.11 | 3 | c.242G>A : p.R81H | rs914917859 |
|  |  | DKFZP434E1119 |  | 11q14.1 | 1 | c.126delC : p.R42fs |  |
|  |  | FUT5 | NM_002034 | 19p13.3 | 5 | c.1046C>A : p.A349D | rs200309409 |
|  |  | HTT | NM_002111 | 4p16.3 | 1 | c.61_63del : p.21_21del | rs1265161631 |
|  |  | IGFN1 | NM_001164586 | 1q32.1 | 12 | c.6114delG : p.L2038fs | not registered,<br>but many<br>variants are<br>registered at<br>this position |
|  |  | MTMR1 | NM_003828 | Xq28 | 1 | c.11_13del : p.4_5del | rs782326539 |
|  |  | NFE2L3 | NM_004289 | 7p15.2 | 4 | c.974A>G : p.N325S | rs143091809 |
|  |  | PCDHB14 | NM_018934 | 5q31.3 | 1 | c.1914C>G : p.H638Q | rs373630707 |
|  |  | PSMB9 | NM_002800 | 6p21.32 | 5 | c.467G>A : p.G156D |  |
|  |  | SPRING1 | NM_024738 | 12q24.22 | 1 | c.1A>G : p.M1V | not registered,<br>but<br>c.1A>T is<br>registered as<br>rs136080880 |

